# Integrating large-scale serum metabolomics and *APOE* ε4 genotype status for the non-invasive detection of Alzheimer’s disease in the ADNI cohort

**DOI:** 10.64898/2025.12.29.25343146

**Authors:** Dany Mukesha, Hüseyin Firat, Guillaume Sacco

**Affiliations:** Firalis SA, Huningue, France; Centre Hospitalier Universitaire de Nice, Nice, France; Université Côte d’Azur, INSERM, CNRS, IPMC, Nice, France

## Abstract

**INTRODUCTION:** Accurate Alzheimer’s disease (AD) detection remains challenging and often requires invasive or costly procedures. Blood-based metabolomic signatures offer a promising non-invasive approach. This study aimed to identify a serum metabolite panel and evaluate its performance alone and in combination with *apolipoprotein E* (*APOE*) ε4 genotype status for distinguishing AD from cognitively normal (CN) individuals.

**METHODS:** Baseline data from 594 participants in the Alzheimer’s Disease Neuroimaging Initiative (237 AD, 357 CN) were analyzed. High-resolution serum metabolomics (Biocrates MxP® Quant 500) and *APOE* genotype data were used for LASSO-based feature selection, followed by machine learning model training and evaluation on a held-out test set.

**RESULTS:** A panel of 151 metabolites distinguished AD from CN with high accuracy (test-set AUC=0.90). Adding *APOE* to the panel further improved model performance (AUC=0.91 versus AUC=0.75 for *APOE* alone; *p*<0.001), achieving strong sensitivity (0.92), specificity (0.84), and negative predictive value (0.94). Key predictive metabolites included bile acids, ether-linked phosphatidylcholines, and acylcarnitines, which are associated with pathways related to lipid metabolism, mitochondrial function, and the gut–liver–brain axis.

**CONCLUSION:** Integrating serum metabolomics with *APOE* enables accurate, non-invasive AD detection and offers a scalable screening approach with strong potential to rule out AD in primary care.

**ClinicalTrials.gov Identifier:** NCT00106899 and related ADNI phases.

## 1. Introduction

The global landscape of Alzheimer’s disease (AD) diagnostics is currently undergoing a transformative shift, driven by the increasing clinical precision and expanding diagnostic accessibility. As of 2024 and 2025, the traditional reliance on invasive lumbar punctures for cerebrospinal fluid (CSF) analysis and high-cost positron emission tomography (PET) imaging has been challenged by the emerging high-performance blood-based biomarkers (BBMs) (1–3). This transition is not merely a change in methodology but a fundamental evolution in how the disease is defined, staged, and managed in clinical practice. The 2024 National Institute on Aging-Alzheimer’s Association (NIA-AA) revised criteria have established AD as a biological process, defined by the presence of core biomarkers of amyloid and tau pathology, regardless of the clinical symptomatic state (4), However, this biological focus has sparked significant debate within the scientific community. Critics, such as the International Working Group (IWG) led by Dubois and colleagues in 2024 (5), argue that AD should remain a clinical-biological construct, requiring both biological evidence of pathology and the presence of objective cognitive symptoms for a formal diagnosis. This tension underscores a critical need for biomarkers that do more than just detect protein aggregates. Therefore, there is a demand for signatures that reflect the systemic physiological state and the metabolic disruptions that drive the progression from early amyloidosis to active, symptomatic neurodegeneration.

Metabolomics, the study of low-molecular-weight metabolites in biological systems, offers a uniquely high-resolution window into these complex pathophysiological shifts. Unlike the genome, which provides a static blueprint of risk, the metabolome is dynamic, reflecting the ongoing interactions between genetics, environment, lifestyle, and disease state. In the context of AD, metabolomics has the potential to capture early signatures of mitochondrial dysfunction, oxidative stress, lipid membrane remodeling, and neurotransmitter imbalance, processes that often preceded the widespread accumulation of amyloid plaques and tau tangles. Large-scale epidemiological investigations, including studies using the UK Biobank dataset (n ≈ 274,000), have already demonstrated that blood-based metabolite profiling can predict incident AD with robust accuracy, particularly when signatures of lipoprotein metabolism, branched-chain amino acids, and glucose homeostasis are integrated with demographic and cognitive features (6–8).

Despite these advances, many prior metabolomics studies have been limited by small sample sizes, a lack of standardized quantification platforms, or a failure to fully integrate metabolic data with the most potent genetic risk factor for sporadic AD: the *apolipoprotein E* (*APOE*) ε4 allele. The *APOE* ε4 isoform is central to the AD narrative, as it is known to impair lipid transport, reduce amyloid clearance, and exacerbate neuroinflammatory responses. The current study seeks to address these gaps by leveraging the Alzheimer’s Disease Neuroimaging Initiative (ADNI) cohort, utilizing the Biocrates MxP® Quant 500 platform for absolute quantification of serum metabolites, and applying advanced machine learning to identify a reproducible metabolic signature that distinguishes AD from cognitively normal (CN) individuals at baseline.

## 2. Material and methods

### Study population and ADNI cohort selection

We analyzed data from the Alzheimer’s Disease Neuroimaging Initiative (ADNI), a landmark multicenter longitudinal study designed to develop and validate clinical, imaging, genetic, and biochemical biomarkers for the early detection and tracking of AD. The ADNI project, which began in 2004, has progressed through several phases (ADNI-1, ADNI-GO, ADNI-2, and ADNI-3), recruiting participants across North America. From an initial pool of 2,418 participants, we carefully selected a sub-cohort of 594 individuals who possessed definitive baseline diagnoses and complete serum metabolomic data.

The diagnostic classification for this cohort was rigorous. The AD group (n=237) consisted of patients meeting the 2024 NIA-AA framework criteria for confirmed AD (9,10). These individuals exhibited objective cognitive impairment, typically characterized by memory complaints verified by a study partner, abnormal scores on the Wechsler Memory Scale (logical memory II), and Mini-Mental State Examination (MMSE) scores between 20 and 26. The cognitively normal (CN) control group (n=357) was defined by MMSE scores between 28 and 30, a Clinical Dementia Rating (CDR) Sum of Boxes (CDR-SB) score of 0, and no evidence of significant depression or other confounding psychiatric conditions. Exclusion criteria were strictly applied to ensure the specificity of the metabolic markers; participants with comorbid neurological disorders (such as Parkinson’s disease or stroke), major psychiatric illness (such as schizophrenia or major depressive disorder), or incomplete metabolomic data were removed from the analysis. The demographic profile of all the participants included age, sex, race, and education level are summarized in the Results section.

The study was approved by institutional review boards at all participating sites, and written informed consent was obtained from all participants prior to the sample collection. Data is available through the ADNI repository. Further information regarding the study protocol can be found at *ClinicalTrials.gov* under identifier NCT00106899.

### Metabolomics profiling and technical specifications

Serum samples were analyzed using the Biocrates MxP^®^ Quant 500 kit, a high-resolution mass spectrometry platform that allows for the absolute quantification of a broad spectrum of metabolites (Fig 1). The Quant 500 panel is designed to cover up to 630 metabolites across several major biochemical families. The platform utilizes a combination of liquid chromatography-tandem mass spectrometry (LC-MS/MS) and flow injection analysis (FIA). This dual approach ensures high sensitivity for polar metabolites like amino acids and biogenic amines, while providing robust quantification for complex lipids.

**Fig 1.**
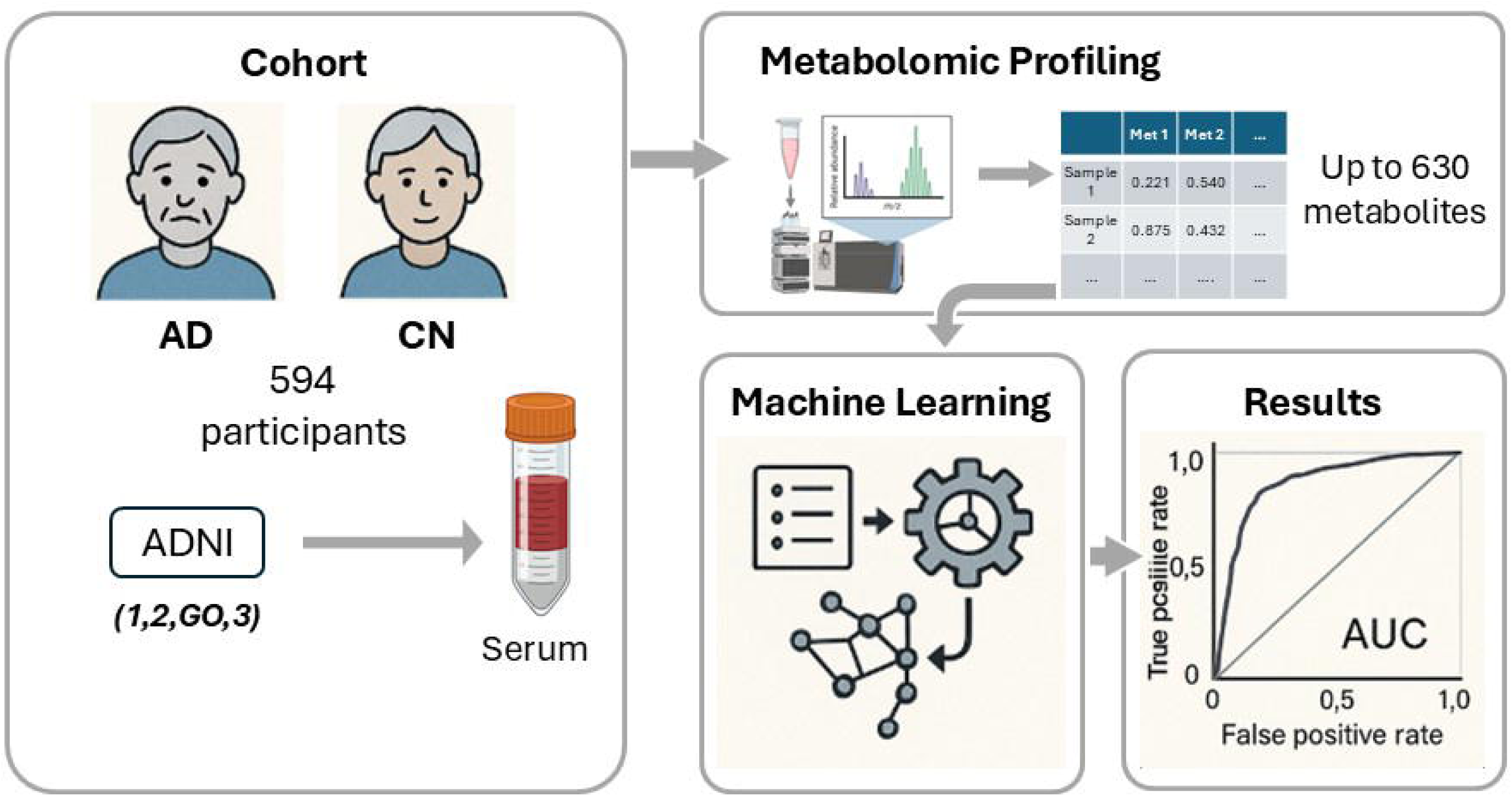
From serum metabolomics patterns to predictive models for accurate detection of AD. The study analyzed metabolomic profiles from 594 participants (AD vs. CN) in the ADNI cohort, measuring up to 630 serum metabolites. Machine learning was applied to develop predictive models, with performance evaluated using AUC (area under the curve) to assess diagnostic accuracy for baseline AD detection. The goal was to derive a highly accurate metabolic signature capable of distinguishing AD from CN individuals at baseline. The workflow highlights the potential of systems-level metabolomics combined with predictive modeling to uncover reproducible biomarkers for AD, addressing the need for non-invasive, scalable screening tools.

Rsigorous data preprocessing and quality control (QC) pipeline was implemented. Initial steps involved the exclusion of metabolites with a high prevalence of missing values (>20%). For the remaining metabolites, missing values were imputed using k-nearest neighbors (k=5), a method that leverages the local structure of the data to provide accurate estimations. Data normalization was achieved via probabilistic quotient normalization (PQN), which corrects for global differences in metabolite concentrations across samples and mitigates batch effects. To ensure technical reliability, any metabolite measurement with a coefficient of variation (CV) exceeding 25% in reference QC samples was discarded. After these quality filtering steps, the final dataset consisted of 623 high-quality metabolites ready for downstream machine learning analysis.

### Feature selection and machine learning pipeline

The analysis aimed to identify the most discriminative metabolic features and build a predictive model optimized for AD detection. The dataset was split into a training set (70% of the cohort, n ≈ 416) and an independent test set (30% of the cohort, n ≈ 178). All feature selection and model tuning were performed exclusively within the training set to prevent optimistic bias and data leakage.

Feature selection was conducted using the least absolute shrinkage and selection operator (LASSO) regression. LASSO is a regularized regression method that adds an L1 penalty to the objective function, effectively shrinking the coefficients of less important variables to zero. This sparsity-inducing property is particularly valuable for metabolomic datasets where high dimensionality and multicollinearity are prevalent. The optimal value for the tuning parameter λ (lambda) was determined using repeated 5-fold cross-validation (Fig S1). Following feature selection, we trained and compared five distinct machine learning algorithms: LASSO, partial least squares (PLS), random forest, extreme gradient boosting (XGBoost), and naive bayes. Models were developed using repeated 5-fold cross-validation (20 repetitions) with stratification by diagnosis and *APOE* status. Potential confounding variables, including age and sex, were adjusted for prior to model construction to ensure that the identified metabolic associations were robustly linked to AD status rather than demographic shifts.

### Performance evaluation and statistical framework

Three feature sets were evaluated: (1) metabolomic data alone, (2) *APOE* ε4 carrier status, and (3) combined metabolomic + *APOE* genotype data. A held-out test set was used for final evaluation. Primary metric for evaluating model performance was the area under the receiver operating characteristic curve (AUC-ROC). The ROC curve plots the true positive rate (sensitivity) against the false positive rate (1-specificity) across various decision thresholds. To ensure the statistical reliability of the performance estimates, 95% confidence intervals (CIs) were calculated using 2000 stratified bootstrap replicates. Secondary metrics, reported at the optimal decision threshold determined by Youden’s index, included sensitivity (Recall), specificity, positive predictive value (PPV), and negative predictive value (NPV). Pairwise comparisons between AUCs were conducted using the DeLong test (11). To further confirm that the models were not overfitted to noise, permutation testing was performed (12,13). This involved shuffling the diagnostic labels 1000 times to build a null distribution of AUC values, against which the actual model performance was compared to establish a p-value. All statistical analyses were performed using R version 4.3.0, the *pROC* package(14) for ROC analyses and the *caret* package(15) for machine learning model development and cross-validation.

## 3. Results

### Demographic and clinical profile

There were no statistically significant differences in age, sex, or race between groups. As expected, cognitive performance measures, including Mini-Mental State Examination (MMSE) and Clinical Dementia Rating Scale-Sum of Boxes) CDR-SB scores, demonstrated clear separation between diagnostic groups. The *APOE* ε4 allele was significantly more prevalent among participants with AD, consistent with established genetic risk patterns. Detailed demographic and clinical characteristics are summarized in Table 1.

**Table 1.**
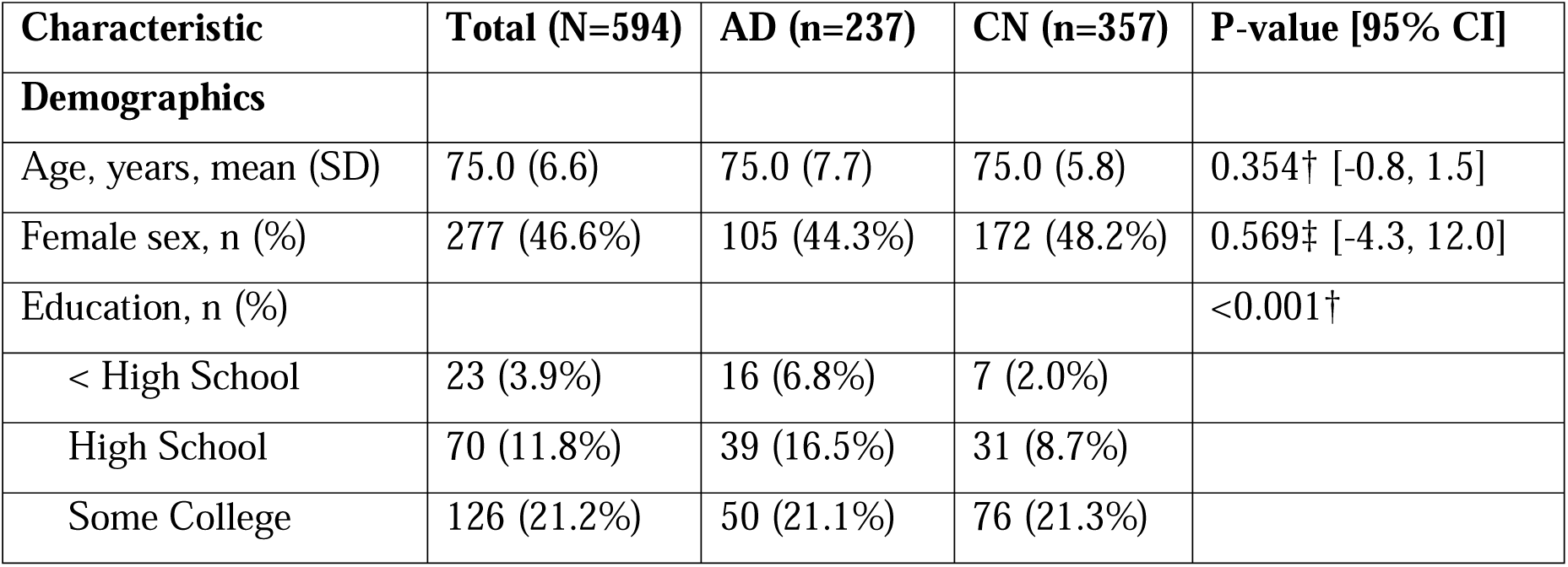
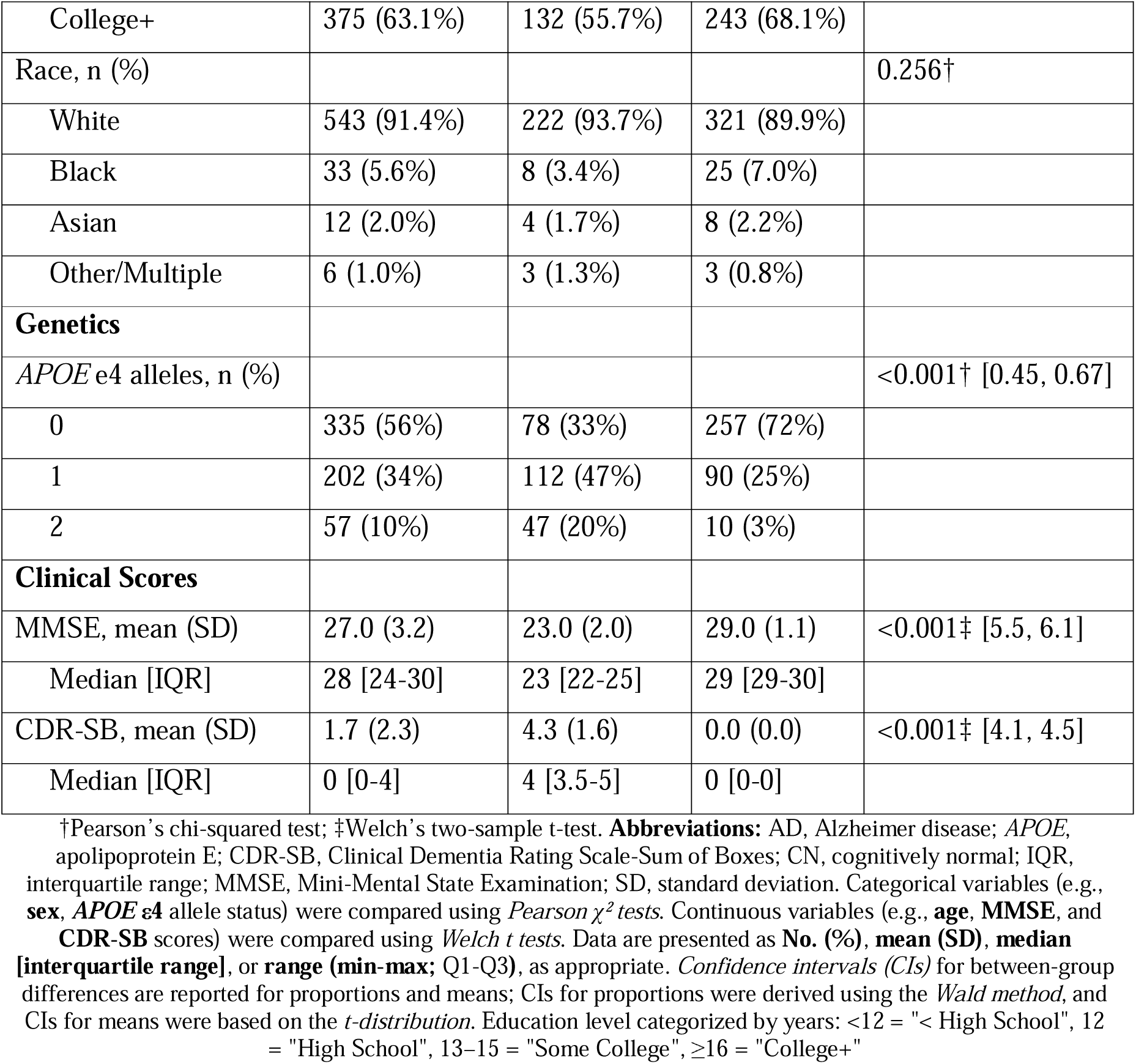
Baseline Characteristics of the Study Cohort.

### Algorithm comparison and model optimization

Evaluation of the five machine learning algorithms on the independent test set revealed that the LASSO and PLS models generally outperformed the other methods in terms of consistency and overall discriminative power (Fig 2A, Table S1). Permutation-based null distributions confirmed that the observed classification accuracy was significantly better than chance (p< 0.001) (Fig 2B). PLS model exhibited a pronounced sensitivity-specificity trade-off, achieving an exceptionally high sensitivity (0.99) but at the cost of lower specificity (0.63). This indicates that while PLS is effective at identifying almost all AD cases, it produces a high number of false positives in cognitively normal individuals. In contrast, the LASSO model provided a more balanced and harmonized performance across all metrics (Table S2).

**Fig 2.**
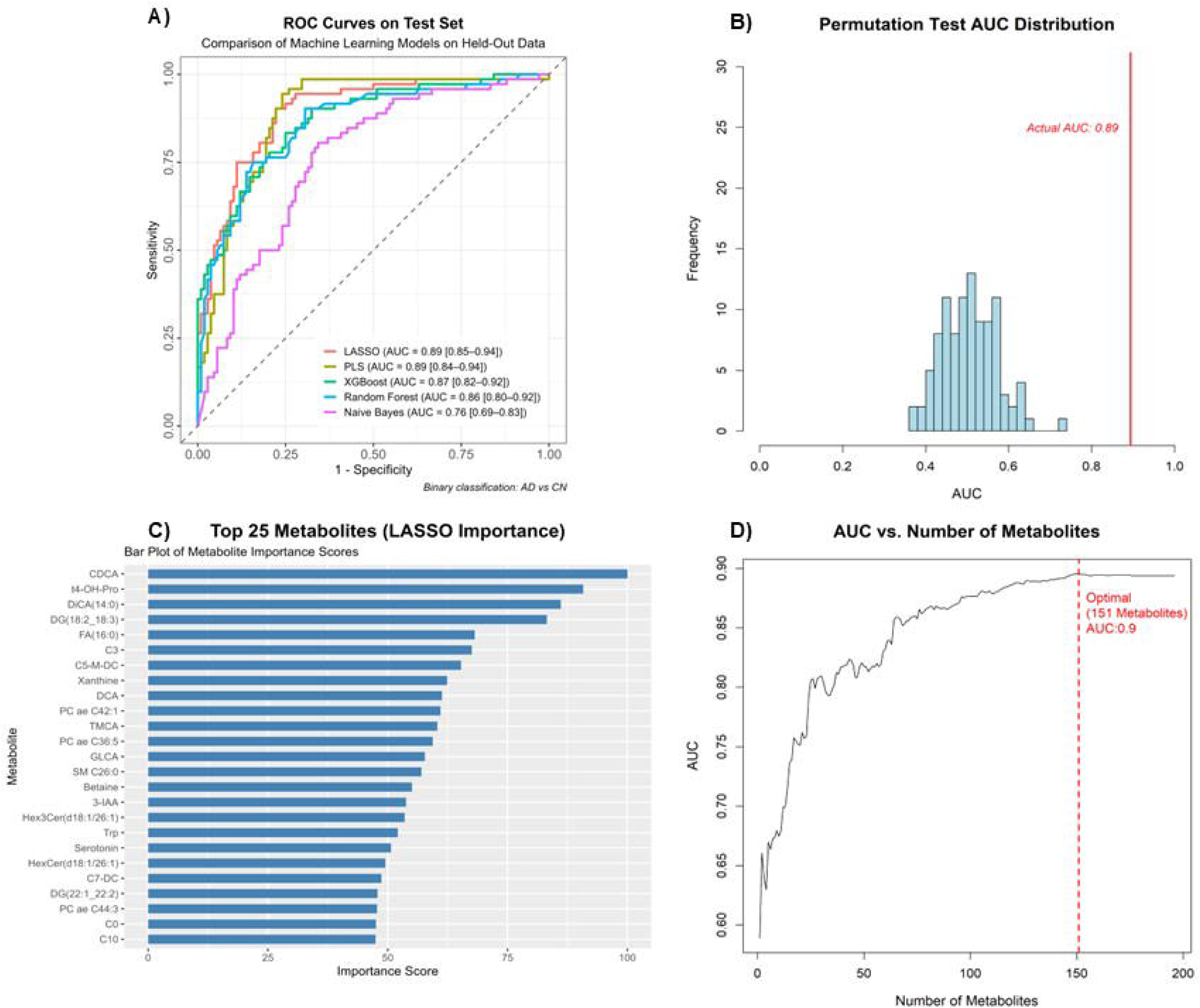
Model performance and metabolite importance in predictive analysis. (A) ROC curves comparing the performance of machine learning models (LASSO, PLS, XGBoost, Random Forest, Naive Bayes) on the test set, with LASSO and PLS showing the highest AUC (0.89). (B) Permutation test with shuffling of training labels only, evaluated on an untouched test set, results confirming the robustness of the LASSO model (actual AUC = 0.89). (C) Bar plot of the top 25 metabolites ranked by importance scores from LASSO, highlighting key biomarkers such as CDCA and t4-OH-Pro. (D) Relationship between the number of metabolites and model performance (AUC), with the optimal subset (151 metabolites) achieving an AUC of 0.9.

Analysis of the relationship between the number of metabolites and model performance indicated a clear plateauing effect (Fig 2D). While initial performance increased rapidly as the first 50 metabolites were added, the gain in AUC became marginal beyond 150 features, which argued against the need for exhaustive metabolite panels, instead pointing to a finite set of mechanistically interlinked analytes. The “Optimal Model” was therefore defined as a LASSO-based classifier utilizing 151 metabolites. This optimized 151-metabolite panel distinguished AD from CN with an AUC of 0.90 (Fig 2D).

Models trained on combined metabolite data and *APOE* genotypes yielded higher AUCs compared to using metabolomics data or *APOE* alone (Fig 3A). This indicates that metabolic profiles provide substantial predictive information beyond genetic risk. Of translational relevance, the incremental predictive value of *APOE* status when combined with metabolomics supported a precision medicine approach where genomic context enhances metabolic risk stratification. The combined model correctly identified 88% of CN and 91% of AD cases, a marked improvement over *APOE*-only predictions (75% CN, 55% AD) (Fig 3B).

**Fig 3.**
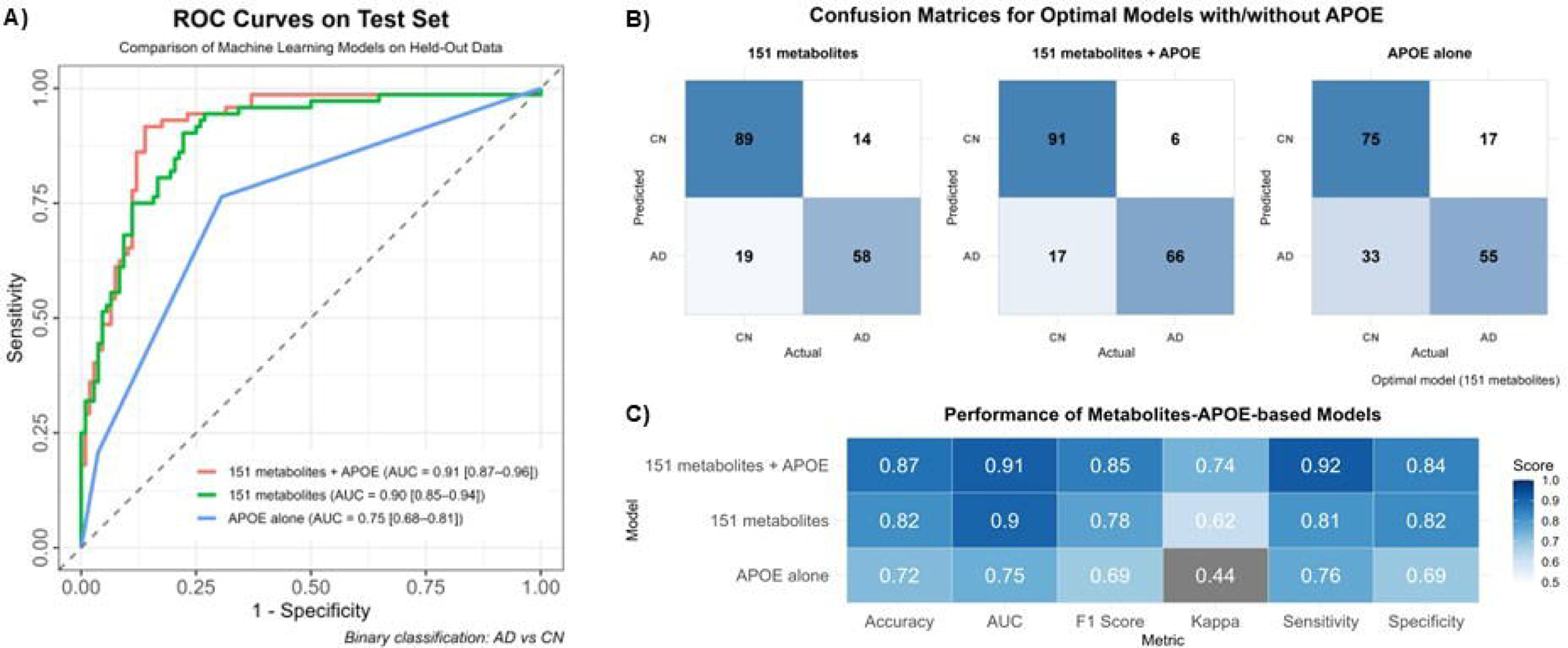
Performance comparison machine learning (ML) models with and without *APOE* in AD Classification. (A) ROC curves comparing LASSO models trained with metabolites only (AUC = 0.90), metabolites + *APOE* (AUC = 0.91), and *APOE* alone (AUC = 0.75) for binary classification of AD vs. cognitively normal (CN) participants. The optimal model (m=151 features) demonstrates improved performance when *APOE* is combined with metabolomic data (B) Confusion matrices and (C) performance metrics (sensitivity, specificity, precision, accuracy) for each model variant. The LASSO model with *APOE* achieves the highest balanced accuracy.

The LASSO model trained on combined features achieved the best results: mean cross-validated AUC of 0.90 (95% CI: 0.88-0.93 with 2000 stratified bootstrap replicates) on top 151 metabolites (Table S1). The final test-set reached AUC of 0.91 (95% CI, 0.87-0.96) with accuracy, recall, precision, and specificity consistently in the high range (Table S2; Fig 3B).

### Key metabolites and biological signature

The top predictor was the primary bile acid chenodeoxycholic acid (CDCA), followed by 4-hydroxyproline (t4-OH-Pro) and tetradecane dioic acid (DiCA(14:0)). Collectively, these three metabolites accounted for >60% of the total feature importance in the model (Fig 2C). While APOE ε4 status alone showed moderate predictive power (AUC = 0.75), its integration with metabolomics significantly improved model performance (ΔAUC = +0.16 vs. *APOE*-only; p<0.001).

The 151-metabolite panel encompassed 22 distinct biochemical classes (Fig S2, Table S3). Lipid species were most represented, with triglycerides, diglycerides, and phosphatidylcholines as dominant subclasses. Several amino acid-related pathways were enriched, including derivatives such as t4-OH-Pro (involved in collagen turnover) and arginine-related metabolites (implicated in nitric oxide signaling), both relevant to vascular contributions to dementia. Multiple sphingolipid subclasses (e.g., ceramides) were identified, aligning with evidence of sphingolipid dysregulation in AD. The high ranking of bile acids underscores the involvement of interconnected pathways spanning energy metabolism, inflammation, and gut-liver-brain axis signaling in AD. Major functional clusters are summarized in Table 2.

**Table 2.**
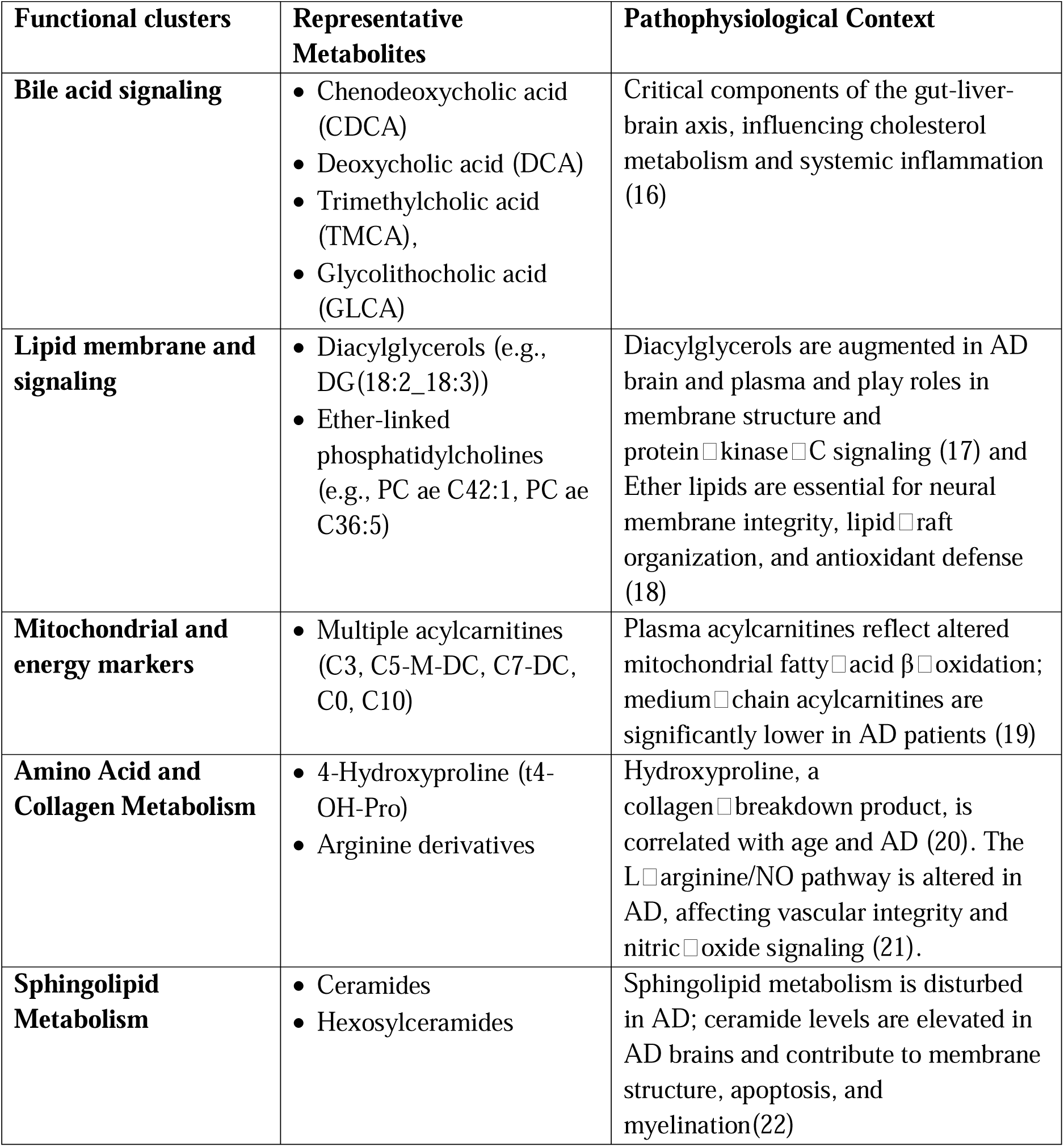
Major functional clusters of identified metabolites.

## 4. Discussion

### Clinical-metabolomic integration and multi-omics synergy

The core insight of this study is that serum metabolomics and *APOE* ε4 status provide synergistic information that enhances AD detection far beyond the capacity of either modality alone. In the independent test set, the integration of *APOE* ε4 into the metabolic model resulted in an absolute improvement in AUC of 0.16 compared to using *APOE* alone (0.91 vs 0.75; Table S2). This synergistic effect supports a precision medicine framework where genomic risk (the “predisposition”) is combined with the metabolic state (the “manifestation”) to refine risk stratification (23–26).

A second-order inference from this synergy is that *APOE* ε4 does not act on AD risk through a single pathway but rather creates a unique metabolic “soil” that is more receptive to pathological processes. The *APOE* ε4 allele is known to impair lipid transport and exacerbate neuroinflammation; the current finding that a lipid-dominated metabolic signature (Fig S2) provides such a strong additive signal suggests that our model is capturing the specific metabolic consequences of being an ε4 carrier. This aligns with recent multi-omics frameworks that position AD as a “network disorder” where genomic susceptibility and downstream metabolic dysfunction (such as lipid peroxidation and mitochondrial stress) co-accelerate neurodegeneration (27).

### Systematic breakdown of lipidomic and membrane pathology

The dominance of lipid species in the prioritized 151-metabolite panel ($n=62$ total lipid species including TGs, DGs, and PCs) provides robust evidence for systemic membrane pathology in AD (28). Phosphatidylcholines are the primary building blocks of neuronal and mitochondrial membranes.8 The presence of numerous ether-linked species (designated with the “ae” prefix in the Biocrates panel, such as PC ae C42:1 and PC ae C36:5) is particularly significant.1 Ether lipids, which include plasmalogens, are highly enriched in lipid rafts and act as critical antioxidants that protect cellular components from oxidative stress. Similar lipid dominance also aligned with post-mortem studies of AD brain tissue but contrasted with association of *APOE* standalone with amyloid pathology, shows that the model captured distinct, potentially co-occurring disease mechanisms (29,30).

A third-order insight here is that the serum metabolic changes we detected likely reflect a global breakdown in lipid raft homeostasis. Lipid rafts are specialized membrane microdomains where the processing of amyloid precursor protein (APP) occurs; disruptions in these domains have been shown to shift APP processing toward the amyloidogenic pathway, increasing the production of Aβ42. The fact that these lipid signatures are detectable in serum indicates that AD pathology involves a systemic failure of membrane integrity that extends beyond the central nervous system, offering a rationale for using blood-based metabolomics as a scalable proxy for brain health.

### The gut-liver-brain axis and peripheral cholesterol homeostasis

The ranking of chenodeoxycholic acid (CDCA) as the most discriminative feature in the panel highlights the importance of the gut-liver-brain axis in AD pathophysiology. Bile acids are synthesized from cholesterol in the liver and are further metabolized by the gut microbiome into secondary bile acids like deoxycholic acid (DCA), which also ranked among the top features ($n=9$).

This connection between bile acids and AD suggests several deep-seated biological trends:

- **Impaired cholesterol clearance:** Altered bile acid profiles in the serum indicate that the peripheral mechanisms for cholesterol disposal are compromised in individuals with AD, potentially leading to the systemic lipid dysregulation captured by the rest of the panel (9).
- **Microbiome involvement**: The inclusion of secondary bile acids and microbial metabolites like indole-3-acetic acid (3-IAA) and hipuric acid points to a disruption in the gut microbiome. Growing evidence suggests that gut-derived metabolites can cross the blood-brain barrier and influence neuroinflammation through the farnesoid X receptor (FXR) and the TGR5 receptor, both of which are expressed in the brain and are sensitive to bile acid signaling.
- **Neuroprotective failures:** Bile acids like CDCA are not just metabolic waste products; they are potent signaling molecules that regulate mitochondrial function and energy homeostasis (16). Their disruption at baseline in the ADNI cohort indicates that these protective signaling pathways are already failing in the early stages of the disease.

### Vascular contributions and extracellular matrix turnover

The identification of 4-hydroxyproline (t4-OH-Pro) as the second most important feature provides a crucial insight into the non-amyloid components of AD. 4-hydroxyproline is a modified amino acid almost exclusively found in collagen; its circulating levels are a direct reflection of collagen turnover and extracellular matrix remodeling.

In the context of AD, this finding likely signals a significant vascular contribution to the disease phenotype. Small vessel disease and the breakdown of the blood-brain barrier (BBB) are nearly universal comorbidities in AD (9). The BBB relies on a robust vascular basement membrane, which is primarily composed of type IV collagen. Elevated or altered levels of t4-OH-Pro in serum may therefore represent the systemic “leakiness” of the vasculature or the degradation of the basement membrane that facilitates the infiltration of neurotoxic peripheral factors into the brain. This captures a facet of “biological aging” and vascular health that is distinct from amyloid-tau-neurodegeneration (ATN) biomarkers, explaining why our model provides complementary information to proteomic tests.

### Mitochondrial exhaustion and acylcarnitine profiles

The presence of 12 distinct acylcarnitines in the 151-metabolite panel (including C3, C5-M-DC, and C10) points to mitochondrial dysfunction as a core component of the AD metabolic signature. Acylcarnitines are essential intermediates in the transport of fatty acids into the mitochondria for β-oxidation. Disruptions in these levels suggest a shift in energy utilization and mitochondrial “exhaustion,” where the brain and peripheral tissues may be failing to efficiently utilize lipids as an alternative fuel source to glucose.

Specifically, the short-chain acylcarnitine C3 is a byproduct of the catabolism of branched-chain amino acids (BCAAs). Altered BCAA metabolism has been repeatedly identified in large cohorts like the UK Biobank as a predictor of incident AD, suggesting that systemic failures in amino acid metabolism and mitochondrial energy production are early events that occur alongside, or even before, clinical symptoms.

### Clinical potential for rule-out screening and population health

The performance of our combined metabolomic-genomic model (AUC = 0.91) is highly competitive with current clinical gold standards, yet it offers significant advantages in terms of scalability and cost. While PET imaging and CSF assays remain the diagnostic definitive tests (31), they are ill-suited for large-scale population screening due to their invasive nature and the requirement for specialized infrastructure.

The high negative predictive value (NPV) of 0.94 is particularly transformative for primary care settings. In a triage scenario, the primary utility of a blood-based biomarker is to “rule out” AD in patients presenting with mild cognitive complaints, thereby avoiding the high cost and potential risks of unnecessary invasive referrals (31,32). A screening tool that can correctly identify 94% of true negatives could drastically optimize healthcare resource allocation while ensuring that high-risk individuals are prioritized for more expensive confirmatory tests.

Furthermore, our model aligns with the July 2025 Palmqvist guidelines (3), which endorse blood-based biomarkers with at least 90% sensitivity and 75% specificity for clinical use. By meeting and exceeding these thresholds (0.92 sensitivity, 0.84 specificity), the 151-metabolite panel integrated with *APOE* represents a regulatory-grade candidate for baseline disease stratification.

### Comparisons with previous large-scale studies

Compared to earlier metabolomics studies, which were often restricted to small “pilot” samples of ∼100 individuals (25), our analysis of 594 participants provides a higher level of statistical power and robustness. The consistency of our AUC (0.91) with findings from the UK Biobank (AUC 0.86–0.87 for incident AD) (33–35) confirms that metabolic signatures provide a stable and reproducible signal across different cohorts and platforms.

Our study also reinforces the specific metabolite classes identified in recent multi-omics ADNI investigations (33–35), particularly the involvement of ether lipids and acylcarnitines. However, by providing absolute quantification via the Biocrates platform, our panel offers a level of standardization and translational relevance that untargeted “discovery-based” metabolomics often lacks. This standardized approach is critical for the development of clinical-grade diagnostic assays that must perform reliably across different laboratories.

### Technical robustness and algorithmic considerations

The superiority of LASSO and PLS algorithms over more complex ensemble methods like Random Forest or XGBoost is a noteworthy technical finding. Metabolomic data is characterized by “wide” datasets (thousands of features, hundreds of samples) and high multicollinearity. The regularized linear approach of LASSO is inherently better suited for this structure because it prevents overfitting by shrinking redundant coefficients to zero, a capability that ensemble methods, which are prone to over-fitting high-dimensional noise, may lack in smaller cohorts.

The plateau in performance at 151 metabolites is another key insight, arguing against the need for exhaustive panels of 600+ metabolites. This indicates that there is a finite, biologically interlinked set of analytes that capture the core metabolic variance of AD. From a clinical development perspective, a focused 151-metabolite panel is significantly easier to validate, manufacture, and clear through regulatory agencies than a larger, less interpretable signature.

### Comparative context for identified blood-based metabolite classes

The broad biochemical spectrum captured by our top-ranked metabolites; spanning lipids, bile acids, and amino acid derivatives; supports prior findings that AD involves complex, multisystem metabolic disturbances. Previous metabolomics studies have similarly reported elevated acylcarnitines and disrupted phospholipid profiles in AD and mild cognitive impairment (MCI) cohorts, though often using narrower panels or untargeted platforms with limited clinical utility (36). In contrast, our panel provides absolute quantification and regulatory-grade standardization, enhancing translational relevance.

Notably, while CSF biomarkers and PET imaging remain clinical gold standards, they are invasive or costly. In comparison, our blood-based metabolite panel integrated with *APOE* genotype achieved classification performance (AUC=0.91) that is competitive with, and in some contexts exceeds, reported accuracies from imaging-based diagnostics (37). This supports the growing rationale for metabolomics-driven precision medicine, where metabolic signatures provide complementary, accessible, and biologically informative tools for early AD risk stratification.

### Evaluation of limitations and generalizability

While the current results are robust, several limitations must be acknowledged. First, the ADNI cohort is predominantly white and North American, which may limit the generalizability of the metabolic signature to more diverse populations where dietary habits, environmental exposures, and genetic backgrounds (beyond *APOE*) are different. Second-order implications of this bias suggest that the specific rankings of metabolites like bile acids, which are heavily influenced by diet and the microbiome, might shift in international cohorts (38). Validation in more representative datasets, such as ARIC or UK Biobank, is essential (39).

In addition, the cross-sectional baseline design of this study means that the signature represents a “snapshot” of established disease rather than a predictor of conversion. Longitudinal studies are required to determine if these 151 metabolites can identify “converters” who are cognitively normal at baseline but will progress to MCI or AD within 2–5 years. Finally, while PQN normalization and rigid QC protocols were used, potential batch effects remain an inherent concern in large-scale multicenter studies like ADNI.

### Future directions: Network-based machine learning and precision prevention

Looking forward, the integration of metabolomics into the AD clinical workflow could be further enhanced by “explainable” machine learning frameworks. One such promising approach is explainable graph-theoretical machine learning (XGML) (40,41). Rather than treating metabolites as independent features, XGML models them as nodes in a metabolic network. By identifying the “edges” or biochemical connections that are most disrupted in AD, we can move from simple diagnostic classification to a deeper understanding of the causal neural-metabolic mechanisms.

Moreover, the synergy between *APOE* ε4 and metabolic signatures opens the door for precision prevention strategies. If ε4 carriers exhibit a specific “lipidomic failure” pattern, targeted nutritional or pharmacological interventions aimed at stabilizing lipid rafts or modulating the gut-liver-brain axis (e.g., through bile acid sequestrants or specific probiotics) could potentially delay the onset of symptoms. This move toward “interventional metabolomics” represents the next frontier in AD research, where biomarkers are used not just for detection but as dynamic guides for therapy (42).

## 5. Conclusion

This study provides a comprehensive and robust validation of a serum-based metabolomic panel for the accurate baseline detection of Alzheimer’s disease. By integrating absolute quantification of 151 prioritized metabolites with *APOE* ε4 genotype status, we have developed a signature that achieves an AUC of 0.91, meeting the rigorous performance standards for clinical triage in specialized memory care settings. The predictive model is characterized by a high negative predictive value (0.94), making it an ideal candidate for rule-out screening in primary care to reduce the burden of unnecessary invasive diagnostics.

The biological insights derived from this study underscore AD as a systemic metabolic disorder, where failures in lipid membrane integrity, gut-liver-brain signaling (bile acids), vascular basement membrane turnover (4-hydroxyproline), and mitochondrial energy production (acylcarnitines) converge to drive pathology. The significant synergy between genetic risk and metabolic manifestation validates the multi-omics approach to neurodegenerative disease stratification. While future validation in diverse, longitudinal cohorts is required to confirm generalizability, these findings represent a critical step toward the goal of providing accessible, accurate, and biologically informative tools for the global management of Alzheimer’s disease.

## Data Availability

Data used in this study were obtained from the Alzheimer's Disease Neuroimaging Initiative (ADNI) database (ClinicalTrials.gov Identifiers: NCT00106899, NCT01078636, NCT01231971, NCT02854033). ADNI is a multicenter longitudinal study, and its data are publicly available to qualified researchers upon application at https://adni.loni.usc.edu/. No new raw data were generated for this study; analyses were performed on existing ADNI data.

## 7. Acknowledgements

The realization of this project was supported and funded by **CombiDiag**, HORIZON – MSCA Doctoral Networks 2021 program under grant agreement (GA): **101071485**. Data collection and sharing for this project were funded by the **Alzheimer’s Disease Neuroimaging Initiative (ADNI)** (National Institutes of Health Grant **U01 AG024904**; Department of Defense award **W81XWH-12-2-0012**).

ADNI is funded by the National Institute on Aging, the National Institute of Biomedical Imaging and Bioengineering, and through generous contributions from the following organizations: AbbVie; Alzheimer’s Association; Alzheimer’s Drug Discovery Foundation; Araclon Biotech; BioClinica, Inc.; Biogen; Bristol-Myers Squibb Company; CereSpir, Inc.; Cogstate; Eisai Inc.; Elan Pharmaceuticals, Inc.; Eli Lilly and Company; EuroImmun; F. Hoffmann-La Roche Ltd and its affiliated company Genentech, Inc.; Fujirebio; GE Healthcare; IXICO Ltd.; Janssen Alzheimer Immunotherapy Research & Development, LLC.; Johnson & Johnson Pharmaceutical Research & Development LLC.; Lumosity; Lundbeck; Merck & Co., Inc.; Meso Scale Diagnostics, LLC.; NeuroRx Research; Neurotrack Technologies; Novartis Pharmaceuticals Corporation; Pfizer Inc.; Piramal Imaging; Servier; Takeda Pharmaceutical Company; and Transition Therapeutics.

The Canadian Institutes of Health Research provides funds to support ADNI clinical sites in Canada. Private sector contributions are facilitated by the Foundation for the National Institutes of Health (www.fnih.org). The grantee organization is the Northern California Institute for Research and Education, and the study is coordinated by the Alzheimer’s Therapeutic Research Institute at the University of Southern California. ADNI data are disseminated by the Laboratory for Neuro Imaging at the University of Southern California.

## 8. Fundings, Conflicts of Interest, and Ethics

## Ethics Approval and Consent

There were no experiments involving animal subjects in this study. Procedures involving human participants were conducted in accordance with the ethical standards of the institutional review boards of the participating ADNI sites and in accordance with the Helsinki Declaration of 1975.

Written informed consent was obtained from all participants by the ADNI study investigators. No identifiable individual data were used in this manuscript.

## Data Availability

The data supporting the findings of this study are available in the public domain from the **Alzheimer’s Disease Neuroimaging Initiative (ADNI)** database. Access is available to qualified researchers upon application at https://adni.loni.usc.edu/.

## Trial registration

Data used in this study were obtained from the Alzheimer’s Disease Neuroimaging Initiative (ADNI) database (ClinicalTrials.gov Identifiers: **NCT00106899**, **NCT01078636**, **NCT01231971**, **NCT02854033**). No new raw data were generated for this study; all analyses were conducted on existing ADNI data.

