## Supplementary Material for "Integrating large-scale serum metabolomics and *APOE* ε4 genotype status for the non-invasive detection of Alzheimer’s disease in the ADNI cohort"

Affiliations:

**Supplementary Figures**

### Fig S1. Machine learning pipeline

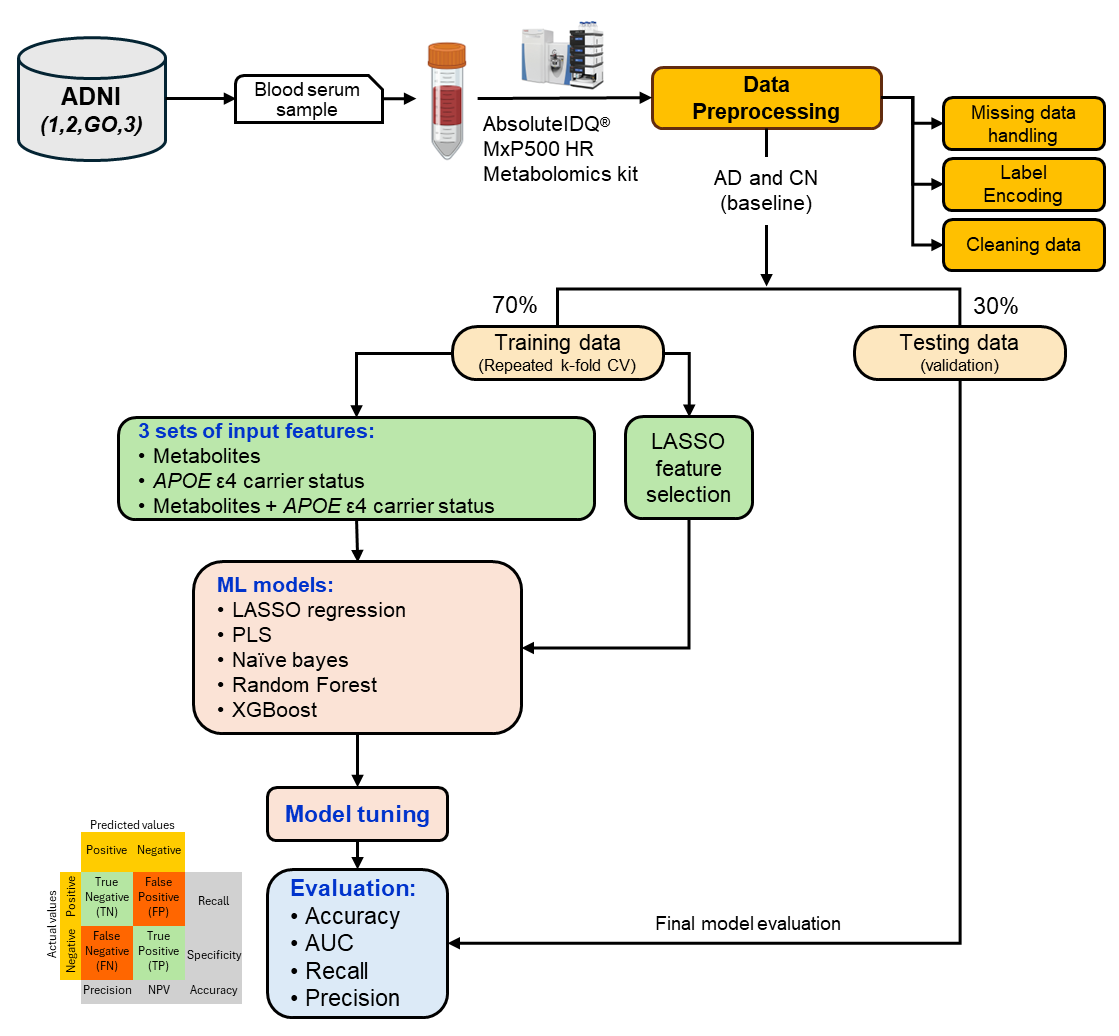

*Description*: This figure illustrates the machine learning (ML) pipeline for AD predictive modeling using ADNI metabolomics data. Blood serum samples from ADNI cohorts (ADNI-1, ADNI-2, ADNI-GO, and ADNI-3) were analyzed with the AbsoluteIDQ® MxP®500 HR kit. After preprocessing (missing data handling, label encoding, and data cleaning), participants were classified as AD or cognitively normal (CN) at baseline. The dataset was split into 70% training (with repeated k-fold cross-validation) and 30% testing. Three feature sets were evaluated: metabolites, *APOE* ε4 carrier status, and their combination. LASSO regression was used for feature selection, followed by testing with multiple ML algorithms (LASSO, PLS, naïve Bayes, random forest, XGBoost). Model tuning was performed on training data, and final evaluation was based on testing set performance, assessed with accuracy, AUC, recall, and precision metrics.

### Fig S2. Biochemical class distribution of 151 metabolites panel

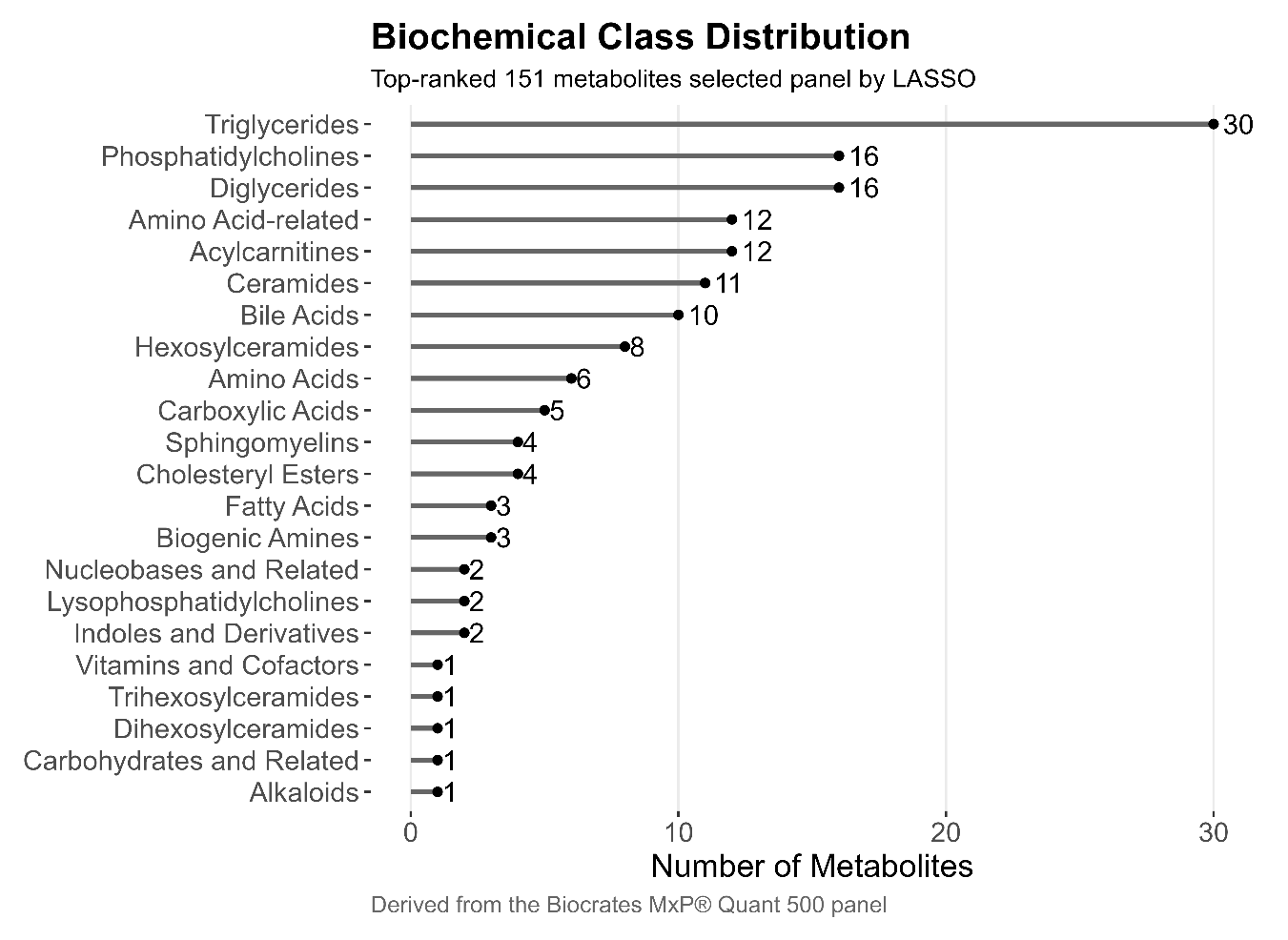
*Description:* Horizontal bar plot showing the biochemical class composition of the top-ranked 151 metabolites selected by Least Absolute Shrinkage and Selection Operator (LASSO) modeling. Metabolites were annotated according to Biocrates MxP® Quant 500 panel classifications and grouped into major biochemical families. Bars represent the number of metabolites per class, with exact counts indicated at the bar termini. The selected feature set is dominated by lipid-related classes, particularly triglycerides (n = 30), phosphatidylcholines (n = 16), and diglycerides (n = 16), followed by amino acid–related metabolites (n = 12), acylcarnitines (n = 12), ceramides (n = 11), and bile acids (n = 10). Additional contributions arise from sphingolipids (e.g., hexosylceramides, sphingomyelins), sterol esters, fatty acids, and smaller numbers of polar metabolites, including amino acids, biogenic amines, nucleobases, vitamins, and carbohydrates. This distribution highlights the prominent involvement of lipid metabolism and associated pathways within the LASSO-selected metabolic signature, while also reflecting a broader, multi-pathway metabolic contribution.

**Supplementary Tables**

### Table S1. Cross-validated performance on training set

| Model | Mean PR AUC | Mean ROC AUC | Lower PR AUC | Lower ROC AUC | Upper PR AUC | Upper ROC AUC |
| --- | --- | --- | --- | --- | --- | --- |
| *196 Metabolite (obtained through Feature selection)* |  |  |  |  |  |  |
| LASSO | 0.89 | 0.84 | 0.88 | 0.83 | 0.89 | 0.85 |
| PLS | 0.89 | 0.84 | 0.89 | 0.84 | 0.9 | 0.86 |
| XGBoost | 0.89 | 0.84 | 0.88 | 0.84 | 0.89 | 0.85 |
| Random Forest | 0.88 | 0.84 | 0.87 | 0.83 | 0.89 | 0.85 |
| Naive Bayes | 0.78 | 0.74 | 0.77 | 0.73 | 0.79 | 0.75 |
| *151 Metabolite*  *(Optimal Model)* |  |  |  |  |  |  |
| LASSO without *APOE* | 0.91 | 0.87 | 0.9 | 0.86 | 0.91 | 0.87 |
| LASSO with *APOE* | 0.92 | 0.89 | 0.91 | 0.88 | 0.93 | 0.9 |
| *APOE genotype alone* |  |  |  |  |  |  |
| LASSO only *APOE* | 0.73 | 0.7 | 0.73 | 0.69 | 0.74 | 0.71 |

*Description:* Performance comparison of five machine learning models using repeated k-fold cross-validation on the training set for AD classification. The mean and 95% confidence intervals (lower and upper bounds) are reported for the area under the precision-recall curve (PR AUC) and the area under the receiver operating characteristic curve (ROC AUC). LASSO, PLS, and XGBoost achieved the highest predictive performance (mean PR AUC = 0.89), while Naive Bayes showed the lowest performance across both metrics. However, Naive Bayes demonstrated the lowest predictive performance across both metrics. When reduced to 151 metabolites (the optimal model), the LASSO model's performance improved further, especially with the inclusion of *APOE* genotype, achieving a mean PR AUC of 0.92. The model using *APOE* genotype alone (LASSO only *APOE*) exhibited the lowest predictive accuracy, with a mean PR AUC of 0.73. All performance estimates derive from internal cross-validation of the training cohort.

### Table S2. External validation metrics on test set

| Model | Accuracy | 95% Confidence Interval | Kappa | Sensitivity | Specificity | Positive Predictive Value (PPV) | Negative Predictive Value (NPV) | F1 Score | Balanced Accuracy | AUC |
| --- | --- | --- | --- | --- | --- | --- | --- | --- | --- | --- |
| *196 Metabolite (obtained through Feature selection)* |  |  |  |  |  |  |  |  |  |  |
| LASSO | 0.82 | (0.75, 0.87) | 0.62 | 0.81 | 0.82 | 0.75 | 0.86 | 0.78 | 0.81 | 0.89 |
| Random Forest | 0.77 | (0.70, 0.83) | 0.53 | 0.76 | 0.78 | 0.70 | 0.83 | 0.73 | 0.77 | 0.86 |
| Naive Bayes | 0.68 | (0.60, 0.75) | 0.33 | 0.58 | 0.74 | 0.60 | 0.73 | 0.59 | 0.66 | 0.76 |
| PLS | 0.77 | (0.70, 0.83) | 0.56 | 0.99 | 0.63 | 0.64 | 0.99 | 0.78 | 0.81 | 0.89 |
| XGBoost | 0.78 | (0.71, 0.84) | 0.55 | 0.78 | 0.78 | 0.70 | 0.84 | 0.74 | 0.78 | 0.87 |
| *151 Metabolite (Optimal Model)* |  |  |  |  |  |  |  |  |  |  |
| LASSO without *APOE* | 0.81 | (0.75, 0.87) | 0.61 | 0.81 | 0.81 | 0.74 | 0.86 | 0.77 | 0.81 | 0.90 |
| LASSO with *APOE* | 0.87 | (0.81, 0.92) | 0.74 | 0.92 | 0.84 | 0.80 | 0.94 | 0.85 | 0.88 | 0.91 |
| *APOE genotype alone* |  |  |  |  |  |  |  |  |  |  |
| LASSO only *APOE* | 0.72 | (0.65, 0.79) | 0.44 | 0.76 | 0.69 | 0.63 | 0.82 | 0.69 | 0.73 | 0.75 |

*Description:* Key classification indices reported for each approach, including accuracy, 95% confidence intervals, Cohen’s kappa, sensitivity, specificity, positive/negative predictive value, F1 score, balanced accuracy, and area under the ROC curve (AUC). Models were trained on sets of selected metabolites (196 or 151 features) and variations incorporating or excluding *APOE* genotype. All performance metrics derive from held-out test set evaluation.

### Table S3. Top 151 metabolites driving best performance in AD vs CN classification.

| n | Metabolite | Class |
| --- | --- | --- |
| 1 | CDCA | Bile Acids |
| 2 | t4-OH-Pro | Amino Acid-related |
| 3 | DiCA(14:0) | Carboxylic Acids |
| 4 | DG(18:2_18:3) | Diglycerides |
| 5 | FA(16:0) | Fatty Acids |
| 6 | C3 | Acylcarnitines |
| 7 | C5-M-DC | Acylcarnitines |
| 8 | Xanthine | Nucleobases and Related |
| 9 | DCA | Bile Acids |
| 10 | PC ae C42:1 | Phosphatidylcholines |
| 11 | TMCA | Bile Acids |
| 12 | PC ae C36:5 | Phosphatidylcholines |
| 13 | GLCA | Bile Acids |
| 14 | SM C26:0 | Sphingomyelins |
| 15 | Betaine | Amino Acid-related |
| 16 | 3-IAA | Indoles and Derivatives |
| 17 | Hex3Cer(d18:1/26:1) | Trihexosylceramides |
| 18 | Trp | Amino Acids |
| 19 | Serotonin | Biogenic Amines |
| 20 | HexCer(d18:1/26:1) | Hexosylceramides |
| 21 | C7-DC | Acylcarnitines |
| 22 | DG(22:1_22:2) | Diglycerides |
| 23 | PC ae C44:3 | Phosphatidylcholines |
| 24 | C0 | Acylcarnitines |
| 25 | C10 | Acylcarnitines |
| 26 | TG(20:3_36:5) | Triglycerides |
| 27 | Cer(d18:0/20:0) | Ceramides |
| 28 | DG-O(18:2_18:2) | Diglycerides |
| 29 | Cer(d18:0/16:0) | Ceramides |
| 30 | DOPA | Amino Acid-related |
| 31 | Pro | Amino Acids |
| 32 | PC aa C34:1 | Phosphatidylcholines |
| 33 | Cys | Amino Acids |
| 34 | DG(18:1_22:5) | Diglycerides |
| 35 | DG(18:1_18:1) | Diglycerides |
| 36 | C16-OH | Acylcarnitines |
| 37 | CE(20:4) | Cholesteryl Esters |
| 38 | DG(18:1_18:4) | Diglycerides |
| 39 | DG(14:0_14:0) | Diglycerides |
| 40 | Spermine | Biogenic Amines |
| 41 | PEA | Biogenic Amines |
| 42 | Trigonelline | Alkaloids |
| 43 | HipAcid | Carboxylic Acids |
| 44 | HexCer(d18:1/24:0) | Hexosylceramides |
| 45 | TG(20:4_36:3) | Triglycerides |
| 46 | PC aa C36:2 | Phosphatidylcholines |
| 47 | SM C16:1 | Sphingomyelins |
| 48 | Sarcosine | Amino Acid-related |
| 49 | TrpBetaine | Amino Acid-related |
| 50 | HexCer(d18:1/18:0) | Hexosylceramides |
| 51 | TG(18:2_36:3) | Triglycerides |
| 52 | DG(18:2_20:0) | Diglycerides |
| 53 | Cer(d18:1/20:0) | Ceramides |
| 54 | PC ae C42:5 | Phosphatidylcholines |
| 55 | TG(16:1_34:3) | Triglycerides |
| 56 | DHA | Fatty Acids |
| 57 | TG(22:6_32:0) | Triglycerides |
| 58 | C9 | Acylcarnitines |
| 59 | TG(16:1_36:2) | Triglycerides |
| 60 | TG(16:0_32:0) | Triglycerides |
| 61 | DG(21:0_22:6) | Diglycerides |
| 62 | Cer(d16:1/20:0) | Ceramides |
| 63 | Anserine | Amino Acid-related |
| 64 | DG(16:0_16:1) | Diglycerides |
| 65 | TG(22:2_32:4) | Triglycerides |
| 66 | PC ae C30:2 | Phosphatidylcholines |
| 67 | TG(16:0_28:2) | Triglycerides |
| 68 | Cer(d18:1/18:0) | Ceramides |
| 69 | TCDCA | Bile Acids |
| 70 | TG(16:1_32:0) | Triglycerides |
| 71 | TG(18:1_32:3) | Triglycerides |
| 72 | Ac-Orn | Amino Acid-related |
| 73 | GDCA | Bile Acids |
| 74 | TG(16:0_34:0) | Triglycerides |
| 75 | AbsAcid | Carboxylic Acids |
| 76 | TLCA | Bile Acids |
| 77 | TG(16:0_38:1) | Triglycerides |
| 78 | DG(17:0_17:1) | Diglycerides |
| 79 | HexCer(d18:1/26:0) | Hexosylceramides |
| 80 | Hex2Cer(d18:1/20:0) | Dihexosylceramides |
| 81 | HexCer(d18:2/20:0) | Hexosylceramides |
| 82 | Choline | Vitamins and Cofactors |
| 83 | TG(18:1_30:2) | Triglycerides |
| 84 | PC aa C38:3 | Phosphatidylcholines |
| 85 | CE(22:6) | Cholesteryl Esters |
| 86 | c4-OH-Pro | Amino Acid-related |
| 87 | lysoPC a C28:0 | Lysophosphatidylcholines |
| 88 | CE(22:5) | Cholesteryl Esters |
| 89 | TG(18:3_35:2) | Triglycerides |
| 90 | CE(16:0) | Cholesteryl Esters |
| 91 | BABA | Amino Acid-related |
| 92 | TG(18:2_36:2) | Triglycerides |
| 93 | Hypoxanthine | Nucleobases and Related |
| 94 | TG(20:3_36:3) | Triglycerides |
| 95 | C12 | Acylcarnitines |
| 96 | H1 | Carbohydrates and Related |
| 97 | C3-DC (C4-OH) | Acylcarnitines |
| 98 | PC aa C42:5 | Phosphatidylcholines |
| 99 | Cer(d18:2/14:0) | Ceramides |
| 100 | TG(18:1_36:3) | Triglycerides |
| 101 | TG(22:4_32:0) | Triglycerides |
| 102 | TG(16:0_34:1) | Triglycerides |
| 103 | TG(18:1_30:1) | Triglycerides |
| 104 | Glu | Amino Acids |
| 105 | HArg | Amino Acid-related |
| 106 | TG(22:5_32:0) | Triglycerides |
| 107 | DG(14:0_18:1) | Diglycerides |
| 108 | PC ae C42:0 | Phosphatidylcholines |
| 109 | TG(20:5_36:3) | Triglycerides |
| 110 | Cer(d18:1/22:0) | Ceramides |
| 111 | HexCer(d16:1/22:0) | Hexosylceramides |
| 112 | beta-Ala | Amino Acid-related |
| 113 | PC ae C38:5 | Phosphatidylcholines |
| 114 | FA(18:0) | Fatty Acids |
| 115 | TG(18:1_38:6) | Triglycerides |
| 116 | C14:1 | Acylcarnitines |
| 117 | TG(17:1_38:7) | Triglycerides |
| 118 | Nitro-Tyr | Amino Acid-related |
| 119 | HexCer(d18:2/22:0) | Hexosylceramides |
| 120 | 3-IPA | Indoles and Derivatives |
| 121 | PC aa C38:4 | Phosphatidylcholines |
| 122 | DG(18:1_20:4) | Diglycerides |
| 123 | PC aa C42:4 | Phosphatidylcholines |
| 124 | Cer(d18:1/20:0(OH)) | Ceramides |
| 125 | PC ae C38:0 | Phosphatidylcholines |
| 126 | DG-O(16:0_20:4) | Diglycerides |
| 127 | TG(20:1_34:0) | Triglycerides |
| 128 | TG(18:1_36:2) | Triglycerides |
| 129 | PC aa C40:3 | Phosphatidylcholines |
| 130 | GLCAS | Bile Acids |
| 131 | DG(16:1_18:2) | Diglycerides |
| 132 | lysoPC a C24:0 | Lysophosphatidylcholines |
| 133 | SM C18:1 | Sphingomyelins |
| 134 | GUDCA | Bile Acids |
| 135 | OH-GlutAcid | Carboxylic Acids |
| 136 | C14 | Acylcarnitines |
| 137 | Cer(d16:1/22:0) | Ceramides |
| 138 | TG(18:2_32:0) | Triglycerides |
| 139 | TG(16:1_36:3) | Triglycerides |
| 140 | C5-OH (C3-DC-M) | Acylcarnitines |
| 141 | TG(18:1_34:4) | Triglycerides |
| 142 | PC aa C36:3 | Phosphatidylcholines |
| 143 | Cer(d18:2/16:0) | Ceramides |
| 144 | Lac | Carboxylic Acids |
| 145 | DG(18:1_22:6) | Diglycerides |
| 146 | CA | Bile Acids |
| 147 | Cer(d18:0/18:0) | Ceramides |
| 148 | Val | Amino Acids |
| 149 | Phe | Amino Acids |
| 150 | SM (OH) C24:1 | Sphingomyelins |
| 151 | HexCer(d18:2/23:0) | Hexosylceramides |

*Description*: List of the top 151 metabolites selected by LASSO modeling, with biochemical class annotations.
